# Impact of COVID-19 Vaccinations in India - A Statewise Analysis

**DOI:** 10.1101/2022.12.02.22283013

**Authors:** Abhigayan Adhikary, Manoranjan Pal, Raju Maiti, Palash Ghosh

**Affiliations:** Economic Research Unit, Indian Statistical Institute, Kolkata, India; Department of Mathematics, Jyoti and Bhupat Mehta School of Health Sciences and Technology, Indian Institute of Technology Guwahati, Assam, India; Centre for Quantitative Medicine, Duke-NUS Medical School, National University of Singapore, Singapore

**Keywords:** COVID-19, Disease modelling, Generalized Gompertz Curves, Forecasting, Time Series, Vaccinations

## Abstract

**Objective:** The COVID-19 vaccination program in India started after the first wave of infections had almost subsided. In this work, the objective is to perform a statewise analysis to assess the impact of vaccination during the second COVID-19 wave in India. A total of 21 states are chosen for the analysis encompassing 97% of the Indian population.

**Methods:** We use the generalized Gompertz curve to study the COVID-19 outbreak. The generalized Gompertz model is then modified to study the impact of vaccination. The modified model considers the cumulative daily number of individuals having the first and second shots of the vaccine in each state as explanatory variables.

**Results:** We observe that, out of 21 states, 16 states show the effectiveness of vaccines in curbing the spread of COVID-19. However, in states like Telangana, West Bengal, Tamil Nadu, Rajasthan, and Kerala, we do not conclusively observe the impact of vaccination during the study period.

**Conclusions:** The effectiveness of COVID-19 vaccine depends on many factors. Some of them are not directly measurable. Using only COVID-19 infection cases and the vaccination data, we conclude that overall the vaccination program was effective in curbing the spread of COVID-19 in India.

## 1 Introduction

Ever since the emergence of COVID-19 and its consequent spread across continents engulfing both advanced and developing nations, there has been a voluminous amount of literature on various aspects of COVID-19. Although a sizable proportion of these contemporary studies highlight the possible robust estimation techniques for modelling COVID-19 infections, the question of assessing vaccine efficacy is yet to be studied rigorously at the *“macro level”* [1]. The existing literature on COVID-19 indicates that the Logistic model and the ARIMA models have been the two popular choices among researchers [2, 3, 4]. The objective of this paper is to perform a statewise analysis to assess the impact of vaccination during the second COVID-19 wave in India.

In this context, [5] used exploratory data analysis to report the COVID-19 situation from January to March 2020 and used the ARIMA model to predict future trends. They predicted a huge surge in COVID-19 infected cases in April and May 2020. They forecasted approximately 7000 COVID-19 infected cases on average in a total span of 30 days in April 2020. However, in reality, the figures were much higher. Similar studies have been performed by [6] and [7] on India, [8] for Pakistan, and [9] for a comparative study of India, Bangladesh, and Pakistan. [10] take a unique approach to forecasting COVID-19 infections for India. They consider the statewise data of infections and model them using the logistic and exponential curves. They infer that the predictions from one model might be misleading and hence suggest a linear combination of the exponential and logistic curves for realistic predictions.

In order to understand the vaccine-induced immunity responses, [11] consider an extension of the classical Kermack–McKendrick model incorporating vaccinations to explore the disease dynamics that last from six months to one year. They infer that vaccine response and its induced immunity are strongly related to the mitigation prevalence. However, the vaccine-induced immunity period remains poorly understood and they validated their claim using the data on COVID-19 deaths in Mexico city and Mexico state. However, unambiguously they observed that natural and vaccine-induced immunity play an essential role in reducing COVID-19 mortality. [12] analyzes EU countries by estimating a non-stationary dynamic panel exhibiting the dynamics of confirmed deaths, infections, and vaccinations per million population from January to July 2021. The study infers that vaccinations alone would not be enough to curb the current and next waves of the COVID-19 pandemic in EU countries. Thus, it becomes evident that the debate on vaccine efficacy is far from being resolved.

[13] has provided some insights on the vaccination status of India through an exploratory data analysis till April 2021. However, the impact of COVID-19 vaccination in India is yet to be analyzed rigorously. The present paper attempts to fill this gap. We go on to explore how the Gompertz Curve can be used as a convenient tool to study the COVID-19 outbreak in India and try to evaluate the role of vaccinations in reshaping the time path of COVID-19 infections in India.

An empirically relevant question that needs to be addressed at this juncture is why not use the Logistic Model intead? Well, it is to be noted that the Gompertz Curves is asymmetric with respect to the point of inflexion whereas the Logistic is symmetric. [14] suggests that when we desire to fit growth curve which show a point of inflection in the early part of the growth cycle, when approximately 35% to 40% of the total growth has been realized, we may use the Gompertz curve with the expectation that the approximation to the data will be good. For COVID-19, the growth rate of infections was much higher in the initial phases as compared to the later phases. Hence, the inflection point of the cumulative infections is expected to occur before the half-life of the wave. So, Gompertz appears to be a more relevant choice.

India experienced a massive crisis in the first wave of COVID-19, and when the nation was contemplating its future strategies, the relatively new and even more deadly “delta variant” became the watchword for the nation since its first detection in November 2020. Although its presence remained mostly insignificant till the end of February 2021, the India-level data shows that soon after the vaccination programme started on 16 January 2021, the number of variants with significant presence grew in number. A wide range of variants showed up in good proportions by early March 2021. By the end of March, the delta variant became *“fitter than others”* and started pushing out the other variants. In the end of May 2021, the delta had a presence of 95%.

However, from the economic viewpoint, the heterogeneity in the way COVID-19 has diffused across the states might render an iota of doubt on the correctness of the results if the analysis is conducted based on the India-level data. Most of the Indian states are large in geographic area and population. Considering the entirety of India to be on the same page, may not provide us with the right picture as suggested by [10]. This is because the new infection rate, preventive measures taken by state governments, and the pace at which vaccinations were carried out are different for each of the states. Hence, there arises a need to analyze the states separately. Thus, we select 21 Indian states for this analysis based on population, which accounts for 97% of the total Indian population during 2020. Figure 1 provides the list of the chosen states.

**Figure 1:**
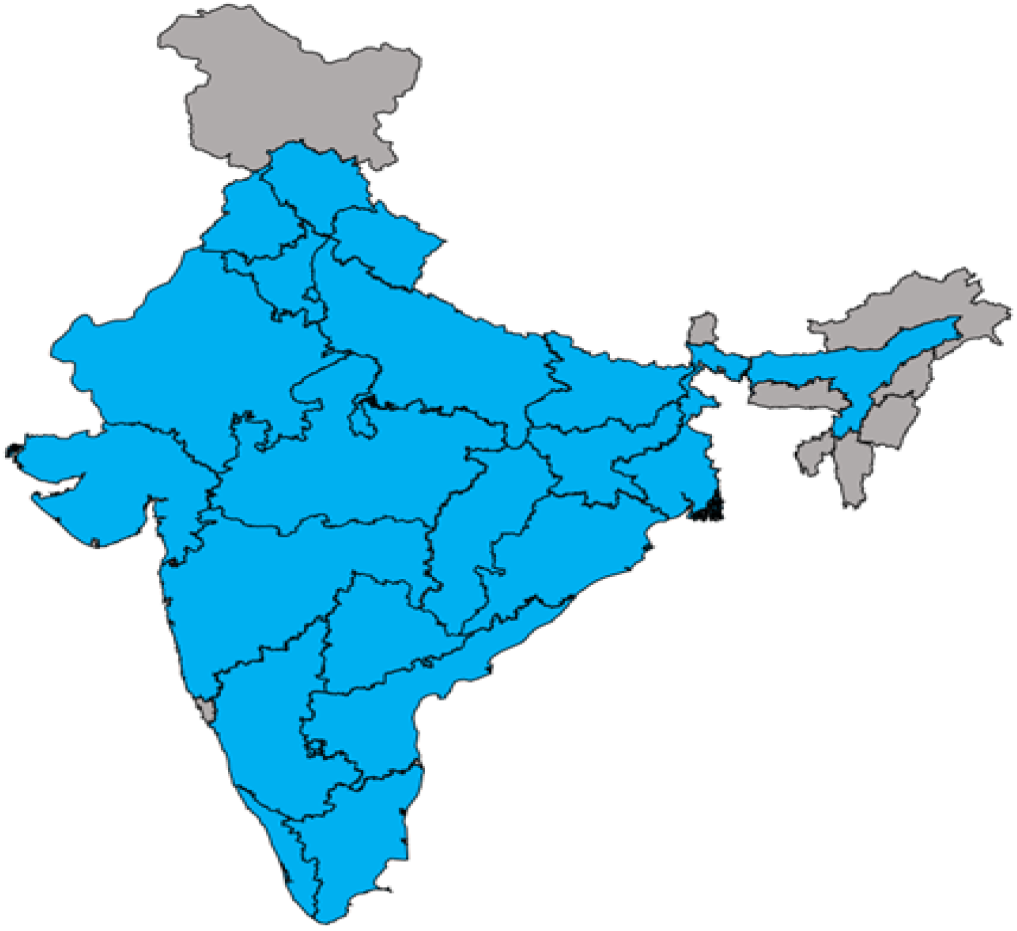
The highlighted states are included in the study

In light of the above background, the paper is divided into the following sections. Section 2 elaborates on the methods used in this paper, followed by the data analysis covered in Section 3. Section 4 highlights our key findings and Section 5 renders the conclusion of the paper.

## 2 Methods

### 2.1 The Gompertz Curves

In recent times, Gompertz Curves have gained popularity for modelling economic and biological phenomena and particularly epidemic modelling. However, only handful of papers have adapted this for their analysis. [15] used the Gompertz Curve to model the COVID-19 cases in USA and also applied it to the data on COVID-19 deaths. They inferred that in both cases, the Gompertz Curve was able to provide a reasonably good approximation to the data. [16] also provided a similar conclusion from their analysis of Italy, Spain, and Cuba. They considered the Logistic curve and the Gompertz Curve and showed that the Gompertz model had better estimates for the peak in confirmed cases and deaths for both the countries. [17] made a detailed comparison of epidemic curves for Sweden and Norway using the Gompertz curves, and they also observed that the epidemic curves for COVID-19 related deaths for most countries with a reliable reporting system are surprisingly well described by the Gompertz-growth model or the Gompertz Curve (GC) given by

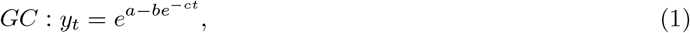

where *a, b, c >* 0. Throughout this paper, we denote *y*_*t*_ as the daily cumulative COVID-19 cases and *N* as the maximum cumulative frequency. From equation (1) we have *N* = *e*^*a*^ which is the asymptote. Note that b is the displacement on the x-axis, and c is the growth rate. Essentially, b and c are the shape parameters.

Equation (1) can also be extended to construct a generalized version of the Gompertz Curve or the Generalized Gompertz Curve (GGC) given by,

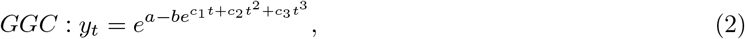

where *a, b >* 0. For the GGC also, *N* = *e*^*a*^ provided *c*_3_ *<* 0. Subsequently, we will also go on to a further extension to incorporate vaccinations in the Gompertz Curve.

### 2.2 The Vaccination augmented Gompertz Curves

Vaccination has been understood as the most prominent external intervention in curbing COVID-19. However, as newer variants of the virus are emerging and leading to new waves of infections, the question of how effective the vaccines are for the Indians is yet to be answered rigorously. Thus, this work develops a framework to assess how vaccinations have reshaped the COVID-19 trajectory in India.

To build a vaccination augmented Gompertz curve, we first fit the GC and the GGC models for each of the states using the data on the second wave of COVID-19 and compare the two. From Table 4, it is clear that the GGC model outperforms the GC model for all the states with respect to 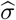, where 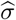 denotes the residual standard error from the fitted regression. Note that only the 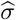 values are reported as our conclusions are unchanged under AIC comparision. Hence, we incorporate the impact of vaccination into the GGC model only.

Consider the following notations and definitions and then a viable extension of the GGC is suggested. Let *t* denote the time unit in days. Define *N*_1*t*_ as the cumulative number of individuals having the first dose and *N*_2*t*_ be the cumulative number of individuals having the second dose at time *t* (in days). Further, define *X*_*t*_ as the cumulative proportion of individuals with the first dose, and *Z*_*t*_ as the cumulative proportion of the individuals with the second dose. Then 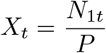 and 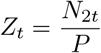, where *P* denotes the population (assumed to be constant during the study period) of the state. In this work, it is sufficient to consider only *X*_*t*_ (because here we want to analyze how an external intervention has modulated one’s ability to resist COVID-19). So, clearly *X*_*t*_ can also be referred to as indicator of the level of vaccinations at any point in time (*t*). It is expected that vaccinations will have a lagged effect, so let *h* denote the lag length in days. We now consider the following extensions of the GGC:

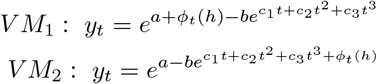

where *ϕ*_t_ (*h*) = *δ*_1_*X* + *δ*_2_*tX* captures the impact of vaccinations, *V M*_1_ and *V M*_2_ denote the two different models that incorporate the effect of the vaccine in different ways. We refer to *ϕ*_*t*_(*h*) as the vaccination function.

Thus, to introduce the impact of external intervention in the form of vaccination, we have considered the above two models. Our motivation behind suggesting this modification can be summarized as follows: one possibility is the introduction of vaccination is likely to reduce the maximum number of cumulative infections (as indicated by *N* = *e*^*a*^) and thereby lowering the asymptote of the Gompertz curve which is the modification considered in *V M*_1_ (as indicated by the modified first exponent term: *a* + *ϕ*_*t*_(*h*)). Also, another possibility is vaccination has a significant contribution in reducing the pace (i.e., the growth rate given by 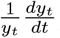) at which the infections occur which we have captured in *V M*_2_ and thereby, the double exponent term has been modified to *c*_1_*t* + *c*_2_*t*^2^ + *c*_3_*t*^3^ + *ϕ*_*t*_(*h*). Hence, for both *V M*_1_ and *V M*_2_, a decreasing *ϕ*_*t*_(*h*) overtime would indicate that vaccines have indeed been effective. However, it is important to note that we might not necessarily have a well-defined asymptote for *V M*_1_. Hence, one cannot proceed with *V M*_1_ for the purpose of modeling. Hence, our appropriate choice for incorporating the effect of vaccinations is *V M*_2_, which shall henceforth be referred to as the Vaccination-augmented Generalized Gompertz Curve (VGGC). Thus, we have:

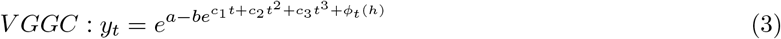

An appealing feature of the formulation presented in Equation (3) is that it enables us to evaluate if there exists a critical level of vaccination say, *X*, beyond which vaccination will be observed to be significantly effective in each of the states, where *t*_0_ indicates the time point at which this value is achieved. In other words, we are willing to begin with the assumption that there exists a critical level of cumulative vaccination beyond which the effect of vaccinations will be remarkably observable in a state and thereby hinting towards a possibility that 100% vaccinations need not be a necessary target to be fulfilled to guarantee the effectiveness of vaccines.

## 3 Data analysis

Data on vaccinated individuals is available from 16 January 2021 to 9 August 2021. Data on the cumulative number of cases is available till 31 October 2021. Both datasets were accessed on 29 January 2022.

### 3.1 Exploratory data analysis of the Vaccination Program

India started its vaccination program on 16 January, 2021 with Covishield and Covaxin initiating the vaccination drive in the country. Note that this was the time when the first wave had nearly subsided in most of the states and the second wave was yet to begin. Thus, the analysis on vaccinations is only relevant for the second wave (often referred as the *delta wave*). The best possible statewise data that is available is on the cumulative number of individuals having the first and the second dose respectively for each of the states (daily data is available till 9 August 2021). Hence, the standard techniques used for assessing the efficacy of vaccines in clinical trials cannot be applied to this aggregate level data simply because an appropriate control group is not available. Hence, evaluating the impact of the vaccination program at the macro level needs further considerations.

The roll out of the vaccination program in India was done in a staggered manner with only the senior citizens (60 years and above) being eligible for getting vaccinated 16 January, 2021 onwards. All individuals 18 years and above became eligible for a shot from 1 May, 2021. As expected, the pace of the vaccination program escalated sharply only after all adults were incorporated in the program. As discussed in Section 2.2, one possibility is that the introduction of vaccination is likely to reduce the maximum number of cumulative infections. This however cannot be directly observed due to the absence of a proper control group. The other possibility is that vaccination has a significant contribution in reducing the pace (i.e. the growth rate) at which the infections occur. This can be analyzed provided certain adjustments are made to account for the different time points at which the second wave started in the different states.

To have some further insights, we will now look at the population adjusted daily new infections 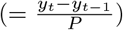 and the population adjusted cumulative vaccinations 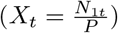 post the beginning of vaccinations for different states. To refrain from further heterogeneities, we will no^*P*^w look at the states with comparable populations like Kerala and Assam, each accounting for 2.604% and 2.598% of the Indian population, respectively.

Figure 2 presents a comparative analysis of Kerala and Assam from 21 April 2021 (the subjectively determined starting point of the Second Wave in Kerala). Note that till this date, the cumulative proportion of the first dose stood at 15.44% and 4.08% respectively. Thus, vaccinations have been quite high in Kerala as compared to that of Assam. Hence, it is natural to expect that the growth of infections will be lower for Kerala or at least will gradually become lower overtime relative to Assam. However, as evident from Figure 2 this is not the case. The new reported infections show a drop but seems to be bouncing back swiftly and one cannot attribute this observation to a slowdown of the vaccination program in Kerala because the cumulative proportion of vaccinations has been consistently higher in Kerala at all time points for the period under consideration as observed from the second panel of Figure 2.

**Figure 2:**
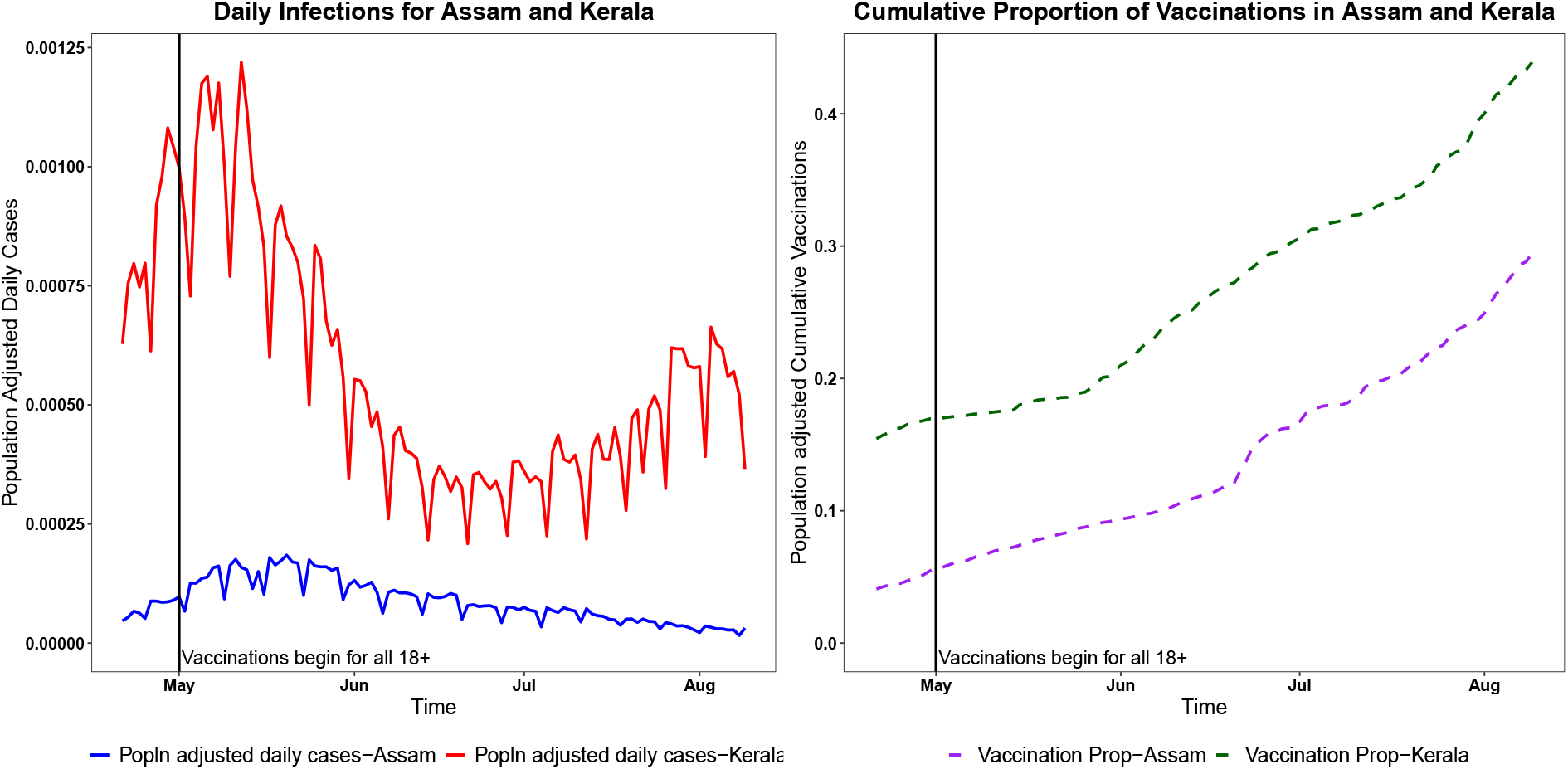
Daily new infections and Vaccinations for Assam and Kerala

An iota of doubt might linger around the observations from Figure 2 owing to the different geographical attributes of Kerala and Assam. To account for this, another comparative study has been done between Telangana and Kerala, each accounting for 2.8% and 2.604% of the Indian population with the cumulative proportion of First dose standing at 7.7% and 15.44% respectively as on 21 April 2021. With Kerala being consistently higher than Telangana with regard to both cases and vaccinations, Figure 3 further confirms the fact that higher vaccination rates has not necessarily translated into lower infection rates for India. This means that we cannot unambiguously claim a cent percent efficiency of the vaccines in controlling the spread of COVID-19.

**Figure 3:**
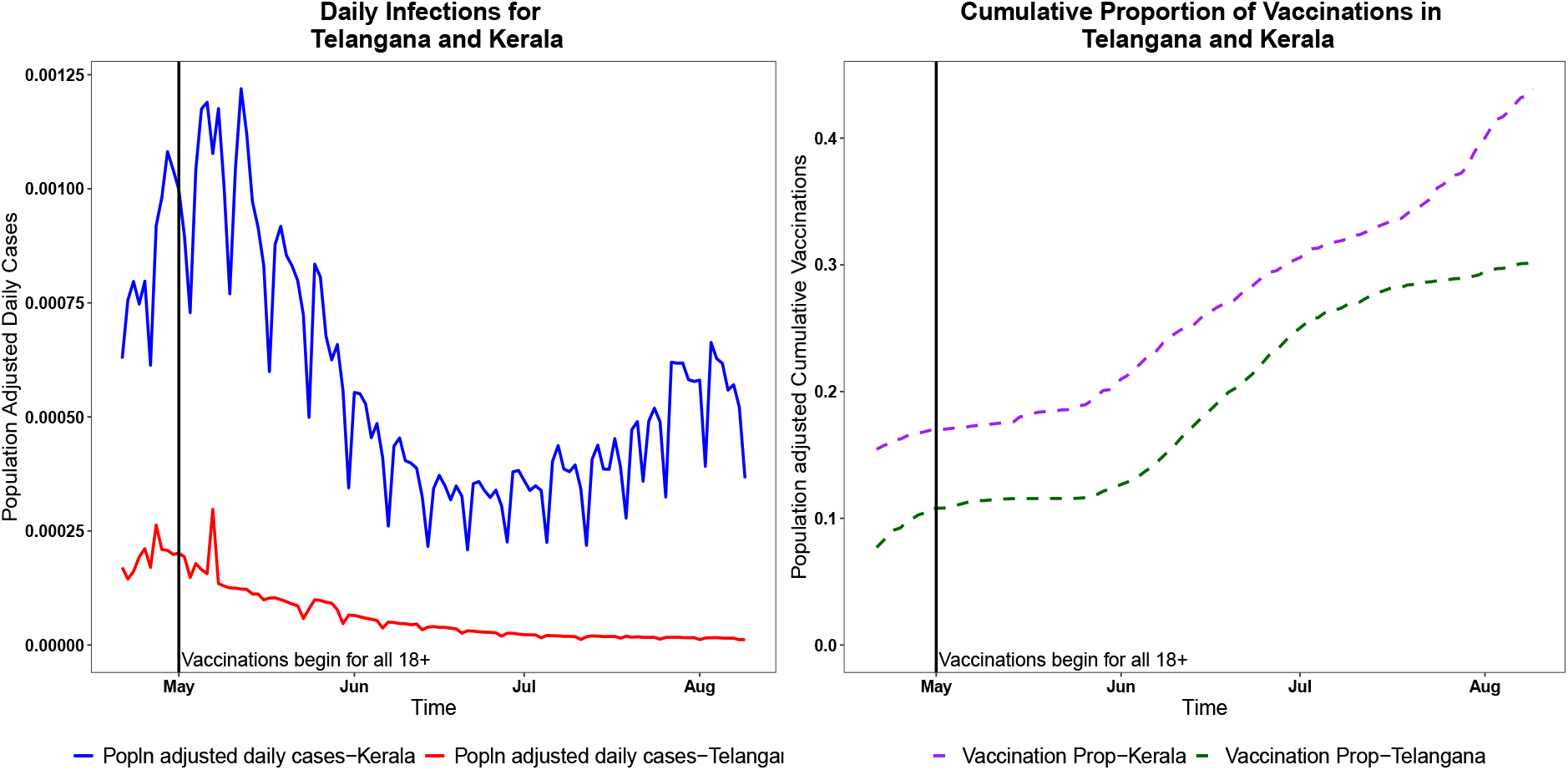
Daily new infections and Vaccinations for Telangana and Kerala

## 4 Results

### 4.1 Determination of the Optimal Lag *h*

Having formulated the VGGC model to study the impact of vaccinations, we turn to a more important question, i.e., what is the optimal value of lag *h* after which the effect of vaccinations is actually observable? To find that we fitted the VGGC model and calculated the vaccination function *ϕ*_*t*_(*h*) by varying the lag *h* between 10 and 50, assuming that the minimum cooling period to get the vaccination effect visible is 10 and maximum is 50. We did it for each state with a grid lengh 10, chosen on an ad hoc basis, i.e., we computed *ϕ*_*t*_(*h*) for *h* = 10, 20, …, 50.

As discussed in Section 2.2, a decreasing *ϕ*_*t*_(*h*) over time would indicate that vaccines have indeed been effective. Hence, for every value of *h*, the number of states displaying this characteristic has been reported in Table 1. The lag corresponding to which we get the highest number of states with a decreasing *ϕ*_*t*_(*h*) will be the optimal which in our case has been observed to be 20. Hence, at the overall level, the optimal lag turns out to be 20 with 16 out of the 21 states exhibiting positive impact of vaccinations. As an illustration of the nature of *ϕ*_*t*_(*h*) for *h* = 20, we provide two different scenarios for states Madhya Pradesh and West Bengal in Figure 4 and 5, respectively. As we can see, in case of Madhya Pradesh, after a certain crtical timepoint (say *t*_0_), *ϕ*_*t*_(20) starts decreasing, whereas for West Bengal there does not exist any *t*_0_ as *ϕ*_*t*_(20) never decreases. In fact, it increases with *t*.

**Table 1:** Lag Lengths(in days) and the number of States(out of 21)

| **Lag Length($h$)** | **Number of states showing a decreasing $\phi_t(h)$** |
| --- | --- |
| **10** | 11 |
| **20** | 16 |
| **30** | 13 |
| **40** | 7 |
| **50** | 3 |

**Figure 4:**
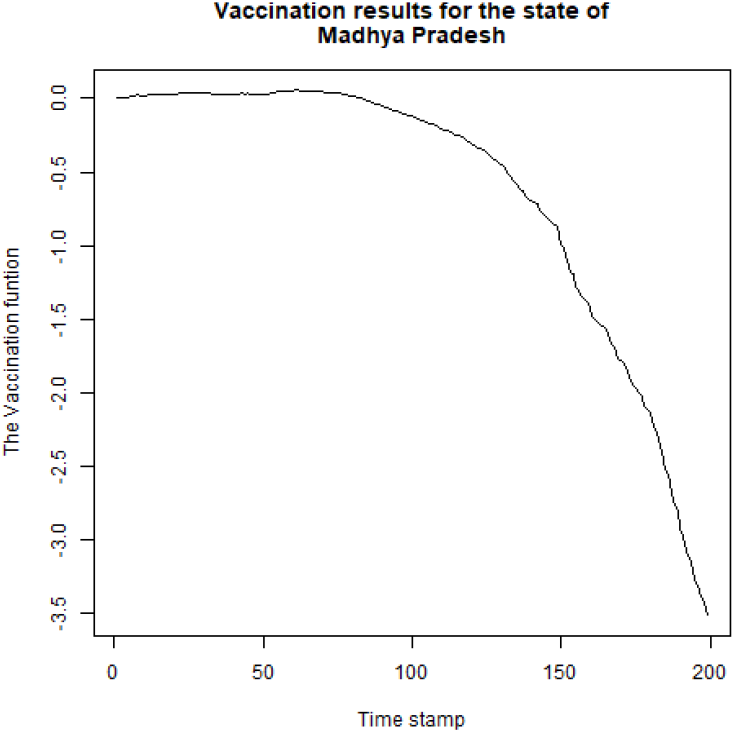
Plot of *ϕ*_*t*_(20) vs t for Madhya Pradesh

**Figure 5:**
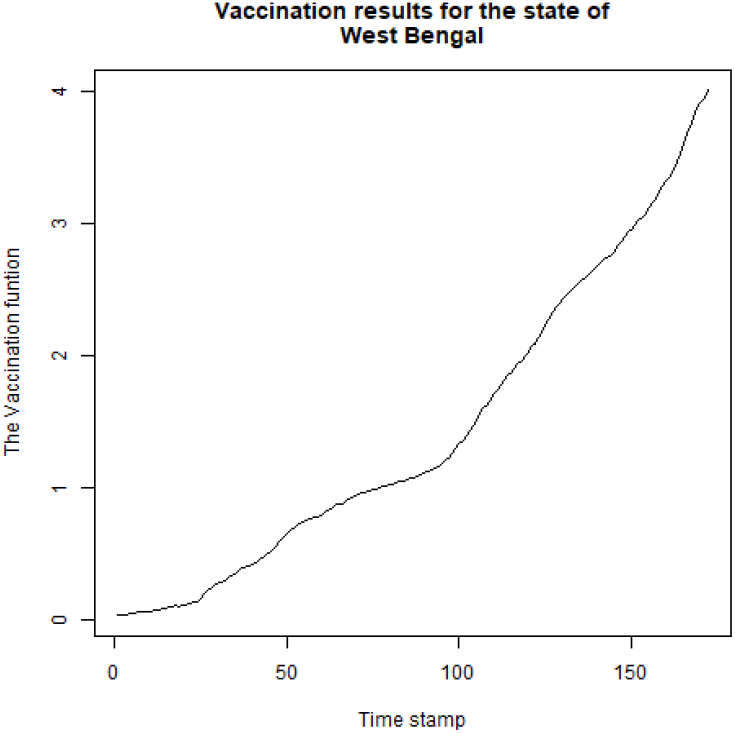
Plot of *ϕ*_*t*_(20) vs t for West Bengal

From the above discussion, it is also evident that a decreasing *ϕ*_*t*_(*h*) must be associated with obtaining a *t*_0_. Hence corresponding to this *t*_0_, we can now obtain a 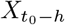 for *h* = 20 (as discussed in Section 2.2). Let us denote this as *X* _20_ which can now be interpreted as the critical level of vaccination after which we observe a decreasing *ϕ*_*t*_ (*h*), i.e, after 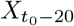, visible positive impact of vaccinations is observable in that particular state. For each of the 16 states exhibiting a decreasing *ϕ*_*t*_ (*h*), the corresponding 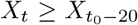 are summarized in Table 2. Note that since the pace of vaccination process differs across the states, the *X*_*T*_ values have also been incorporated in Table 2, where *X*_*T*_ indicates the cumulative proportion of vaccination that has been completed as of 9 August 2021 (the last timepoint in our dataset). As already mentioned, it is observed that a decreasing *ϕ*_*t*_(*h*) was not found for 5 states out of 21 states, namely: Telangana, West Bengal, Tamil Nadu, Rajasthan, Kerala and thus 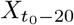 values are also not computable for these states. In Section 4.2 we have used the optimal value of *h* as 20 for our forecasting results. It is important to note that although with the data at hand, a *t*_0_ and a corresponding 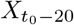 was not observed for the 5 states, if more data is available, these states can be re-examined for this possibility.

**Table 2:** Estimated 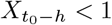 *for all states for lag=20*.

| States | Cumulative vaccinations ($X_T$) | $X_{t_0-20}$ |
| --- | --- | --- |
| Telangana | 30.25 | — |
| Assam | 29.48 | 2.66 |
| Jharkhand | 21.72 | 6.56 |
| Bihar | 18.71 | 5.02 |
| Madhya Pradesh | 34.82 | 8.5 |
| Himachal Pradesh | 57.03 | 13.95 |
| Gujarat | 44.00 | 10.81 |
| Chhattisgarh | 31.19 | 15.5 |
| West Bengal | 23.35 | — |
| Odisha | 30.10 | 11.34 |
| Uttarakhand | 45.07 | 11.79 |
| Andhra Pradesh | 32.70 | 22.48 |
| Karnataka | 38.26 | 9.22 |
| Maharashtra | 28.46 | 0.02 |
| Punjab | 26.56 | 7.12 |
| Tamil Nadu | 26.77 | — |
| Haryana | 35.76 | 7.18 |
| Uttar Pradesh | 19.31 | 4.93 |
| Rajasthan | 33.33 | — |
| Delhi | 41.88 | 15.77 |
| Kerala | 43.90 | — |

### 4.2 Prediction performance of the VGGC Model over the GGC model

In this section, we examined the forecasting performance of the VGGC model over the GGC model to assess the effectiveness of the inclusion of the vaccination program in the GGC model. For each state, we divided the data into the taining set and test set. Since the data on vaccinations is available till 9 August 2021, our training dataset for each state includes cumulative infections from the subjectively determined cutoff point to 18 August 2021, and the test dataset is from 19 August to 28 August 2021. Test set contains the last 10 observations to study the short term future prediction of the model. To facilitate the model comparison, we used the predicted mean squared error (PMSE) for the test set given by

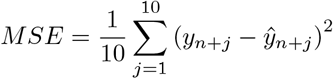

where *y*_*n*+*j*_ denotes the *j*-th test sample observation and *n* is the size of the training dataset. The model giving the lower test MSE will be better in terms of prediction. The results are summarized in Table 3. Note that in Table 3, “Choice=1” indicates that the VGGC performs better than GGC and “Choice=0” indicates otherway round.

**Table 3:** 10 days prediction comparision for the two models (19th August-28th August)

| States | MSE(GGC) | MSE(VGGC) | Choice |
| --- | --- | --- | --- |
| Telangana | 1807.63 | 239.77 | 1 |
| Assam | 736.63 | 1940.35 | 0 |
| Jharkhand | 1369.24 | 1222.63 | 1 |
| Bihar | 2327.26 | 1626.53 | 1 |
| Madhya Pradesh | 1767.74 | 1679.33 | 1 |
| Himachal Pradesh | 8074.99 | 5747.32 | 1 |
| Gujarat | 2249.47 | 1459.29 | 1 |
| Chhattisgarh | 5662.96 | 3596.28 | 1 |
| West Bengal | 16573.27 | 16246.99 | 1 |
| Odisha | 8749.36 | 7771.73 | 1 |
| Uttarakhand | 1968.76 | 1316.84 | 1 |
| Andhra Pradesh | 31516.36 | 30401.4 | 1 |
| Karnataka | 38141.35 | 39246.84 | 0 |
| Maharashtra | 164319.1 | 105972.1 | 1 |
| Punjab | 22228.6 | 1722.45 | 1 |
| Tamil Nadu | 48744.2 | 22016.11 | 1 |
| Haryana | 2002.17 | 1760.22 | 1 |
| Uttar Pradesh | 2516.02 | 2580.59 | 0 |
| Rajasthan | 1343.03 | 1530.53 | 0 |
| Delhi | 3462.38 | 3817.16 | 0 |
| Kerala | 49609.69 | 42491.37 | 1 |

**Table 4:** Fitted Models for the Second Wave

| State | $\hat{\sigma}_{GC}$ | $\hat{\sigma}_{GGC}$ |
| --- | --- | --- |
| Telangana | 8433 | 2150 |
| Assam | 3515 | 3085 |
| Jharkhand | 3155 | 1030 |
| Bihar | 4614 | 1532 |
| Madhya Pradesh | 12440 | 2277 |
| Himachal Pradesh | 3899 | 2025 |
| Gujarat | 14850 | 2667 |
| Chhattisgarh | 9552 | 3981 |
| West Bengal | 12670 | 5505 |
| Odisha | 5436 | 4779 |
| Uttarakhand | 4386 | 1383 |
| Andhra Pradesh | 10410 | 9589 |
| Karnataka | 25580 | 15840 |
| Maharashtra | 77840 | 50410 |
| Punjab | 16350 | 11240 |
| Tamil Nadu | 35330 | 12340 |
| Haryana | 12590 | 1465 |
| Uttar Pradesh | 77840 | 50410 |
| Rajasthan | 12370 | 1956 |
| Delhi | 12680 | 3670 |
| Kerala | 83030 | 13830 |

Here also we get similar conclusions as earlier, i.e., VGGC fit giving better predictions for 16 of the 21 states with the exception of Assam, Karnataka, Uttar Pradesh, Rajasthan and Delhi. Hence, we can conclude that the vaccination augmented Gompertz Curve has given us significant insight into the impact of vaccinations in India as it gives a higher prediction accuracy for the majority of the states under consideration.

## 5 Conclusion

The current paper takes a sequential approach to facilitate the inclusion of vaccinations in modelling the cumulative number of COVID-19 infections. As observed for the state-level data, the GGC gives a better fit for all the states. Vaccinations seem to be a meaningful inclusion in modelling the daily cumulative infections and the optimal lag has been obtained to be 20 days at the *“macro level”*, i.e the effectiveness of the vaccines can be strongly seen after 20 days of the first dose for 16 of the chosen 21 states. This has been backed by our observations that in these states we have been able to observe a 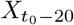. Hence, 100% vaccination is not the gold standard that needs to be necessarily achieved to prove that vaccinations have indeed been effective.

An attempt has been made to statistically answer the question of whether the COVID-19 vaccines are capable of checking infection growth. Our analysis suggests that the claim cannot be fully upheld with certainty at the aggregate level. Although there are a myriad of factors influencing the surge of COVID-19 cases, the fact that 5 states(as obtained in Section 4.1) which are home to 24.28% of Indians not showing visible effects of vaccinations is indeed an observation that cannot be relegated to the background.

There is no gainsaying that one might attribute the counter intuitive conclusions found in the 5 states coming out as a result of some other factor(s) that might dominate the vaccination effect.However, inclusion of these non-medical interventions(like lockdowns, quarantine etc) has not been possible in this analysis due to lack of appropriate and reliable data. Further, the study by [18] on India finds that risk perceptions and social media exposure has nearly insignificant influence on people’s attitudes towards COVID-19 vaccinations. Social norms, trust, and people’s attitudes towards the COVID-19 vaccinations are are the key factors driving their intentions to take up COVID-19 vaccinations. Hence, the issue of vaccine hesitancy cannot be ruled out. Further research needs to be done to address the question of acquired immunity vs natural immunity which continues to remain the fulcrum of assessing vaccine efficacy. The future prospects of this analysis can be focussed on developing improved methodologies to include the other seemingly relevant variables subject to the data availability and thereby robustifying our process of filtering out the vaccination effect on the infections spread.

## Data Availability

The updated data on infections can be downloaded from
\url{www.kaggle.com/datasets/vinitshah0110/covid19-india?resource=download
}, and the statewise data on vaccinations is available at
\url{www.kaggle.com/datasets/harveenchadha/india-covid19-vaccination-data?select=vaccination.csv}

https://www.kaggle.com/datasets/vinitshah0110/covid19-india?resource=download

## Data Availability Statement

The updated data on infections can be downloaded from https://data.covid19india.org/, and the statewise data on vaccinations is available at www.kaggle.com

